# Genotype-phenotype correlations in *SCN8A*-related disorders reveal prognostic and therapeutic implications

**DOI:** 10.1101/2021.03.22.21253711

**Authors:** Katrine M Johannesen, Yuanyuan Liu, Cathrine E Gjerulfsen, Mahmoud Koko, Lukas Sonnenberg, Julian Schubert, Christina D Fenger, Ahmed Eltokhi, Maert Rannap, Nils A. Koch, Stephan Lauxmann, Johanna Krüger, Josua Kegele, Laura Canafoglia, Silvana Franceschetti, Thomas Mayer, Johannes Rebstock, Pia Zacher, Susanne Ruf, Michael Alber, Katalin Sterbova, Petra Lassuthová, Marketa Vlckova, Johannes R Lemke, Ilona Krey, Constanze Heine, Dagmar Wieczorek, Judith Kroell-Seger, Caroline Lund, Karl Martin Klein, PY Billie Au, Jong M Rho, Alice W Ho, Silvia Masnada, Pierangelo Veggiotti, Lucio Giordano, Patrizia Accorsi, Christina E Hoei-Hansen, Pasquale Striano, Federico Zara, Helene Verhelst, Judith S.Verhoeven, Bert van der Zwaag, Aster V. E. Harder, Eva Brilstra, Manuela Pendziwiat, Sebastian Lebon, Maria Vaccarezza, Ngoc Minh Le, Jakob Christensen, Mette U Schmidt-Petersen, Sabine Grønborg, Stephen W Scherer, Jennifer Howe, Walid Fazeli, Katherine B Howell, Richard Leventer, Chloe Stutterd, Sonja Walsh, Marion Gerard, Bénédicte Gerard, Sara Matricardi, Claudia M Bonardi, Stefano Sartori, Andrea Berger, Dorota Hoffman-Zacharska, Massimo Mastrangelo, Francesca Darra, Arve Vøllo, M Mahdi Motazacker, Phillis Lakeman, Mathilde Nizon, Cornelia Betzler, Cecilia Altuzarra, Roseline Caume, Agathe Roubertie, Philippe Gélisse, Carla Marini, Renzo Guerrini, Frederic Bilan, Daniel Tibussek, Margarete Koch-Hogrebe, M Scott Perry, Shoji Ichikawa, Elena Dadali, Artem Sharkov, Irina Mishina, Mikhail Abramov, Ilya Kanivets, Sergey Korostelev, Sergey Kutsev, Karen E Wain, Nancy Eisenhauer, Monisa Wagner, Juliann M Savatt, Karen Müller-Schlüter, Haim Bassan, Artem Borovikov, Marie-Cecile Nassogne, Anne Destrée, An-Sofie Schoonjans, Marije Meuwissen, Marga Buzatu, Anna Jansen, Emmanuel Scalais, Siddharth Srivastava, Wen-Hann Tan, Heather E Olson, Tobias Loddenkemper, Annapurna Poduri, Katherine L Helbig, Ingo Helbig, Mark P Fitzgerald, Ethan M Goldberg, Timo Roser, Ingo Borggraefe, Tobias Brünger, Patrick May, Dennis Lal, Damien Lederer, Guido Rubboli, Gaetan Lesca, Ulrike BS Hedrich, Jan Benda, Elena Gardella, Holger Lerche, Rikke S Møller

## Abstract

We report detailed functional analyses and genotype-phenotype correlations in 433 individuals carrying disease-causing variants in *SCN8A*, encoding the voltage-gated Na^+^ channel Na_V_1.6. Five different clinical subgroups could be identified: 1) Benign familial infantile epilepsy (BFIE) (n=17, normal cognition, treatable seizures), 2) intermediate epilepsy (n=36, mild ID, partially pharmacoresponsive), 3) developmental and epileptic encephalopathy (DEE, n=191, severe ID, majority pharmacoresistant), 4) generalized epilepsy (n=21, mild to moderate ID, frequently with absence seizures), and 5) affected individuals without epilepsy (n=25, mild to moderate ID). Groups 1-3 presented with early-onset (median: four months) focal or multifocal seizures and epileptic discharges, whereas the onset of seizures in group 4 was later (median: 39 months) with generalized epileptic discharges. The epilepsy was not classifiable in 143 individuals. We performed functional studies expressing missense variants in ND7/23 neuroblastoma cells and primary neuronal cultures using recombinant tetrodotoxin insensitive human Na_V_1.6 channels and whole-cell patch clamping. Two variants causing DEE showed a strong gain-of-function (GOF, hyperpolarising shift of steady-state activation, strongly increased neuronal firing rate), and one variant causing BFIE or intermediate epilepsy showed a mild GOF (defective fast inactivation, less increased firing). In contrast, all three variants causing generalized epilepsy induced a loss-of-function (LOF, reduced current amplitudes, depolarising shift of steady-state activation, reduced neuronal firing). Including previous studies, functional effects were known for 165 individuals. All 133 individuals carrying GOF variants had either focal (76, groups 1-3), or unclassifiable epilepsy (37), whereas 32 with LOF variants had either generalized (14), no (11) or unclassifiable (5) epilepsy; only two had DEE. Computational modeling in the GOF group revealed a significant correlation between the severity of the electrophysiological and clinical phenotypes. GOF variant carriers responded significantly better to sodium channel blockers (SCBs) than to other anti-seizure medications, and the same applied for all individuals of groups 1-3.

In conclusion, our data reveal clear genotype-phenotype correlations between age at seizure onset, type of epilepsy and gain- or loss-of-function effects of *SCN8A* variants. Generalized epilepsy with absence seizures is the main epilepsy phenotype of LOF variant carriers and the extent of the electrophysiological dysfunction of the GOF variants is a main determinant of the severity of the clinical phenotype in focal epilepsies. Our pharmacological data indicate that SCBs present a therapeutic treatment option in early onset *SCN8A*-related focal epilepsy.

## Introduction

Since the first pathogenic *SCN8A* variant was discovered in an affected individual with epilepsy^1^, a wide clinical spectrum of neurodevelopmental phenotypes has been reported. The spectrum ranges from benign familial infantile epilepsy (BFIE) with self-limiting seizures and typical cognitive development^2-4^, over an intermediate phenotype with variable seizure onset, treatable seizures and mild intellectual disability (ID)^5^ to early onset developmental and epileptic encephalopathies (DEE) with moderate to severe ID^6-10^, often with movement disorders, cortical visual impairment, severe gastrointestinal symptoms and increased risk of premature death^7, 10-15^. Furthermore, rare clinical presentations with ID, autism spectrum disorder (ASD) and movement disorders without epilepsy have been described^16-19^.

*SCN8A* encodes Na_v_1.6, which is one of four voltage-gated sodium channels expressed in the mammalian brain. Na_v_1.6 is found in the central and peripheral nervous system with a predominant expression in excitatory, but also in inhibitory neurons^20^. Previous studies have revealed that BFIE and DEEs are caused by missense variants with gain-of-function (GOF) effects, whereas truncating variants, deletions and certain missense variants causing loss-of-function (LOF) or both LOF and GOF effects have been associated with ID, ASD and movement disorders with or without seizures^3, 11, 17, 19, 21^.

Treatment of *SCN8A*-related DEEs revealed frequent resistance to anti-seizure medications (ASMs), although treatment with sodium channel blockers (SCBs), especially high-dosage phenytoin, were beneficial in some affected individuals^22^.

Here, we combined a detailed clinical analysis of the largest cohort of individuals with *SCN8A*-related neurodevelopmental disorders investigated to date with functional studies of newly detected variants in mammalian cells and primary neurons and explored the genotype-phenotype correlations in functional studies, computational modeling and treatment response in *SCN8A*.

## Methods

### Cohort ascertainment and phenotyping

Affected individuals were recruited through a network of collaborating clinicians, as well as GeneMatcher^23^, by means of a standardized phenotyping sheet to assess clinical characteristics (medical history, seizure and physical characteristics, family history, neurodevelopment and cognition), EEG, neuroimaging and retrospective data on antiepileptic treatment. Seizures and epilepsy syndromes were classified according to the latest ILAE guidelines^24, 25^.

Based on information on presence and severity of the epilepsy, seizure onset and cognitive status the affected individuals were categorized into the following subgroups:

1. BFIE – infantile onset seizures with offset during infancy/early childhood and normal cognitive development^3^
2. Intermediate epilepsy – individuals with a focal epilepsy of intermediate severity, reflecting neither BFIE nor DEE^5^
3. DEE^14^
4. Generalized epilepsy
5. Unclassifiable epilepsy (insufficient data to be classified into one of the groups above)
6. No epilepsy

Treatment response was evaluated by the referring providers as seizure freedom (at least six months without seizures), seizure reduction (affected individual still on ASMs since provider and parents considered a beneficial effect), no change (ASMs terminated) or seizure aggravation noted by treating providers and parents. Phenytoin (PHT), carbamazepine (CBZ), oxcarbazepine (OXC), lacosamide (LCM), lamotrigine (LTG) and zonisamide (ZNS) were all classified as SCBs. Variant pathogenicity was assessed according to the ACMG guidelines^26^. *SCN8A* transcript NM_014191.3 was used for coding variant nomenclature.

### Previously published cases

A PubMed search on “*SCN8A*” was performed, and all publications with affected individuals were included in the present study. The latest search was performed on May 15^th^ 2020. Papers not available in English were excluded. Only original cases and only probands were included. Data on functional studies were also collected. If affected individuals were published more than once, all papers are included in the list. Duplications of affected individuals were avoided by follow-up with the referring clinician/corresponding author.

### Ethics

The study was approved by the local ethical committees or followed other local guidelines. Previously unpublished individuals (or parents, in case of minors) signed informed consent.

### Data availability

Data will be made available upon request.

### Frequency

The Danish Epilepsy Centre is the only tertiary hospital in Denmark specialized in treating epilepsy, and the majority of affected individuals with non-acquired early onset epilepsy are referred to the center for genetic testing. Furthermore, inquiries were sent to all national clinical genetic departments. Thus, we were able to estimate the frequency of *SCN8A* variants in the Danish population by using the birth cohort from 2006 to 2017 in the electronical population database of the Danish National Statistics.

### Functional studies

All methods have been previously described^19^ and are only briefly summarized in the Supplement.

### Neuronal simulations

To investigate how changes in Na^+^ current properties affect neuronal firing behavior, we simulated a model neuron with sodium, potassium and leak currents (details provided in supplementary methods). We simulated firing by injecting step currents of 0 to 0.75 nA for 2s and analyzed the voltage traces for the firing rate after the neuron adapted to the injected current. The resulting input-output curve was then analyzed for its area under the curve (AUC). We modified the following six parameters of the *Na*^+^ current. The half activation voltages *V*_1/2_ of the activation and inactivation curves were shifted within ±10 mV of the original values. The slopes *k* of the activation and inactivation curves as well as the maximum conductance *g*_Na_ were multiplied with a scaling factor ranging from 0.5 to 2. The persistent current *z* of inactivation ranged from 0 to 5% of the maximum inactivation. Only a single parameter was modified at a time. The effect of alteration *i* on the AUC was quantified relative to the AUC of the unchanged model *AUC*_0_ by

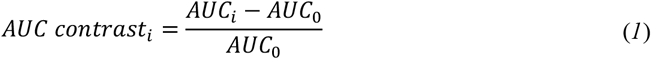

For each altered parameter, the slope *m*_j_ of the regression line through the origin between the parameter changes *x* and its resulting *AUC contrasts* was calculated. To compare magnitudes of effects across different alteration scales, parameter changes *x* were set to Δ*V*_1/2_ = *V*_1/2,i_ − *V*_1/2,0_, Δ*z* = *z*_i_ − *z*_0_ and *log*(*scaling factor*_i_) for *k* and *g*_Na_.

These slopes were then used to score biophysical changes *j* of ionic current variants to estimate their effect on the firing behavior and the severity of their neurological impairments with

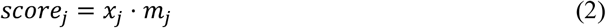

where *score*_j_ is the scoring for one alteration of a biophysical parameter. This resulted in the following equations for the biophysical scores of SCN8A variants:

where *score*_j_ is the scoring for one alteration of a biophysical parameter. This resulted in the following equations for the biophysical scores of SCN8A variants:

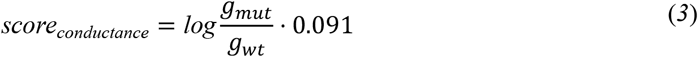

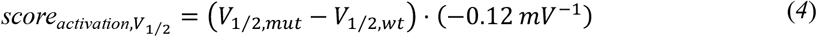

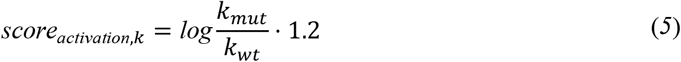

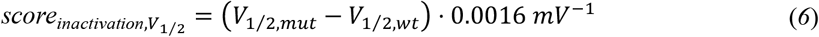

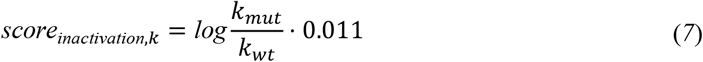

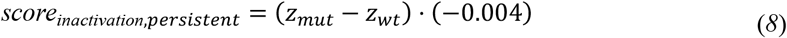

Finally, these scores are summed up to predict the severity of the respective variants

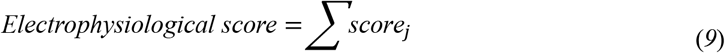

### Classification of phenotype severity

For the correlation analysis between electrophysiological score and severity of the disease in *SCN8A* variant carriers, we classified the individuals in four severity groups: BFIE, IE, DEE with mild or moderate ID, and DEE with severe ID. We assigned weights to the four grades of severity (BFIE=1, IE=2, DEE with mild or moderate ID=3, DEE with severe ID=4). In case that the same variant was associated with different phenotypes, we calculated a weighted average score.

### Statistical analysis

Clinical data was analysed using Stata version 15.1 for Mac (StataCorp, College Station, Texas, USA). For categorical data, Fisher’s Exact Test was used, and for continuous data the Kruskal-Wallis test was used. Significance was evaluated using a two-tailed test of proportions and significance was reached if p<0.05. The do-file used to perform the analyses is available upon request.

Electrophysiological data were analysed using Clampfit software of pClamp 10.6 (Axon Instruments), Microsoft Excel (Microsoft Corporation, Redmond, WA, USA), or Igor Pro (Wavemetrics, Portland, OR, USA). Statistics were performed using one-way ANOVA with Dunnett′s posthoc test or ANOVA on ranks with Dunn′s posthoc test in Graphpad prism (Graphpad software, San Diego, CA, USA). For all statistical tests, significance compared to controls is indicated in the figures using the following symbols: * p < 0.05, ** p < 0.01, *** p < 0.001.

## Results

### Phenotypes within the whole cohort

We assessed a cohort of 95 unpublished and 338 previously published affected individuals (433 affected individuals in total). One hundred and forty-three affected individuals were unclassifiable due to a lack of information and have not been included in the analyses below. Age at follow-up/inclusion ranged from two months to 44 years (mean 4.3 years). We differentiated the following phenotypes:

- BFIE: 17 affected individuals had BFIE. Median age at seizure onset in this subgroup was six months (range: two weeks to 17 months). The most prevalent seizure types were focal (46.2%), focal to bilateral tonic-clonic (38.5%) and bilateral tonic-clonic without identifiable focal onset (38.5%). Cognition was normal in all affected individuals. Treatment response was available in 13 affected individuals, all of whom were seizure free: Seven with an SCB, two with a non-SCB and three with a combination. Nine/17 (52.9%) variants were inherited from an affected parent. Two variants were recurrent: p.(Glu1483Lys) and p.(Asn1877Ser)^2, 3^.
- Intermediate epilepsy: 36 affected individuals had a focal epilepsy fitting neither BFIE nor DEE categories. Median age at seizure onset was five months (range: two months to seven years). The most prevalent seizure types were focal (63.9%), tonic (30.6%), and bilateral tonic-clonic (52.8%). Cognition was normal in 30.6%. Mild ID was present in 55.6% and moderate ID in 13.9%. Additional features included speech delay (36.1%), behavioral disorders (ADHD, autistic features, aggression) (19.4%) or ataxia (11.1%). Treatment response was available in 32 affected individuals; 15 affected individuals were seizure free: eight with an SCB, four with a non-SCB and three with a combination. Three variants were inherited from an affected parent, while seven were unknown and the remainder were *de novo*. Two variants were recurrent: p.(Gly1475Arg) and p.(Asn1877Ser)^5^.
- DEE: 191 affected individuals had a DEE phenotype. Median age at seizure onset was three months (range: first day of life to 36 months). The most frequent seizure types were focal (37.7%), tonic (40.8%) and bilateral tonic-clonic (47.6%) seizures. Cognition ranged from moderate (22%) to severe (69.6%) ID and was unknown in the remainder (8.4%). Additional features included hypotonia in 47.6% and cortical vision impairment (CVI) in 29.3%. Treatment response was known in 128 affected individuals: 26 affected individuals (20.3%) were seizure free, 11 of them with SCBs (42.3%), seven with non-SCBs (26.9%) and eight with a combination (30.8%). The vast majority of variants were *de novo* (89.5%), eight were inherited from an affected parent, in thirteen the inheritance was unknown. Twenty-five variants were recurrent. P.(Arg850Gln/Gly) and p.(Arg1872Trp/Gln/Leu) were the most common^10, 14, 27^.
- Generalized epilepsy: 21 affected individuals had generalized epilepsy. The median age at seizure onset was three years (range: nine months to 14 years). The most prevalent seizure types were absence (81.0%), generalized tonic-clonic (23.8%) and febrile seizures (14.3%). Cognition was normal in 19.0%, and ranged from mild (28.6%) or moderate (38.1%) to severe ID (9.5%). Cognition was unknown in 4.8%. Additional features included ataxia (28.6%), behavioral disorders (autism, aggression, anxiety; 28.6%), and speech delay (19.0%). Treatment response was known in 17 affected individuals; seven were seizure free: two with an SCB and five with a non-SCB. Ten variants occurred *de novo*, five were inherited from an affected parent, and six were of unknown inheritance. None of the variants was recurrent.
- Neurodevelopmental disorder without epilepsy: 25 affected individuals did not have epilepsy at time of inclusion (median age nine years, range three to 35 years). Cognition ranged from normal (8.0%) to mild (36.0%), moderate (24%) or severe ID (12%), and was unknown in 20%. Additional features included behavioral disorders (ASD, ADHD, 36.0%), delayed speech (20.0%) and microcephaly (16.0%). Seven variants occurred *de novo*, while ten were inherited and the remainder was unknown. Three variants were recurrent: p.(Gly384Arg), p.(Arg931Gln) and p.(Ala1622Asp).

Clinical details for all previously unpublished affected individuals and the published affected individuals (as data were available) are summarized in supplementary table 1.

**Table 1.** Clinical characteristics of LOF and GOF variants + phenotypic subgroups

|  | BFIE | Intermediate epilepsy | DEE | Generalized epilepsy | LOF | GOF |
| --- | --- | --- | --- | --- | --- | --- |
| Number of patients | 17 | 36 | 191 | 21 | 32 | 133 |
| Percentage with epilepsy | 100% | 100% | 100% | 100% | 59.4% | 100% |
| Median age at seizure onset | 6 months | 5 months | 3 months | 39 months | 28.5 months | 4 months |
| Most common seizure types | Focal, focal to bilateral TC and bilateral TC | Focal, bilateral TC and tonic | Bilateral TC, focal and tonic | Absences, bilateral TC and febrile seizures | Absence seizures, bilateral TC and myoclonic seizures | Bilateral TCs, tonic and focal |
| Phenotype subgroups | - | - | - | - | Generalized epilepsy 43.8%<br>No epilepsy 34.4%<br>DEE 6.3%<br>Unclassifiable epilepsy 15.6% | BFIE 7.5%<br>Focal epilepsy 10.5%<br>DEE 54.1%<br>Unclassifiable 27.8% |
| Cognition | Normal 100% | Normal 30.6%<br>Mild ID 55.6%<br>Moderate ID 13.9% | Moderate ID 22.0%<br>Severe ID 69.6%<br>Unknown 8.4% | Normal 19.0%<br>Mild ID 28.6%<br>Moderate ID 38.1%<br>Severe ID 9.5%<br>Unknown 4.8% | Normal 15.6%<br>Mild ID 37.5%<br>Moderate ID 18.8%<br>Severe ID 18.8%<br>Unknown 9.4% | Normal 11.3%<br>Mild ID 9.0%<br>Moderate ID 15.0%<br>Severe ID 39.1%<br>Unknown 25.6% |
| Co-morbidities | Paroxysmal kinesigenic dyskinesia | Language delay, behavioral issues | Hypotonia, CVI, ataxia | Language delay, behavioral issues | Language delay, autism, behavioral issues, ataxia | Hypotonia, CVI, dyskinesia, ataxia |
| Mortality | 0% | 0% | 9.4% | 0% | 0% | 9.0% |
| Precision medicine | Sodium-channel blockers | Sodium-channel blockers | Sodium-channel blockers |  |  | Sodium-channel blockers |

Abbreviations: BFIE: Benign familial infantile epilepsy, bilateral TCs: bilateral tonic clonic seizures, CVI: Cortical vision impairment, DEE: Developmental and epileptic encephalopathy, GOF: Gain of function, ID: Intellectual disability, LOF: Loss of function

### Functional studies

We examined seven *SCN8A* variants functionally, chosen to represent the most important aspects of the clinical spectrum, particularly the newly identified phenotype with generalized epilepsy, for which functional studies have not been performed up to now. Three variants (p.(Leu840Pro), p.(Phe846Ser), p.(Asn1877Ser)) were seen in affected individuals with early onset seizures; the first two in affected individuals with DEE^7^, the last in affected individuals with BFIE or intermediate epilepsy^2, 5^. Three variants (p.(Ile1654Asn), p.(Val1758Ala), p.(Thr1787Pro)) were seen in affected individuals with generalized epilepsy with late onset absence seizures, in two cases preceded by febrile seizures^5, 28^. The last variant (p.(Asn374Lys)) was found in an affected individual with a focal epilepsy with moderate ID and late onset at seven years of age^5^.

The biophysical consequences of these seven variants were first studied in ND7/23 cells. P.(Phe846Ser) and p.(Leu840Pro) variants induced hyperpolarising shifts of the activation curves, indicating clear GOF effects (Figs. 1B ,1C, Supplementary Table 2). The p.(Asn1877Ser) variant caused a significant depolarising shift of steady-state fast inactivation and slightly slowed the time course of fast inactivation (Fig. 1D, 1F, Table S2). In contrast, p.(Ile1654Asn), p.(Val1758Ala) and p.(Thr1787Pro) variants dramatically reduced the peak current density; especially cells transfected with p.(Ile1654Asn) barely exhibited any Na^+^ current, hence no gating parameters were obtained for this variant (Figs. 2A, 2B, Supplementary Table 2). Furthermore, the p.(Val1758Ala) and p.(Thr1787Pro) variants shifted activation curves toward more depolarised potentials suggesting LOF effects, although the former variant impaired, and the latter enhanced fast inactivation (Figs. 2B, 2C, 2D, 2F, Supplementary Table 2). Additionally, these two variants, as well as p.(Asn374Lys) to a minor extent, accelerated recovery from fast inactivation (Fig. 2E, Supplementary Table 2).

**Figure 1.**
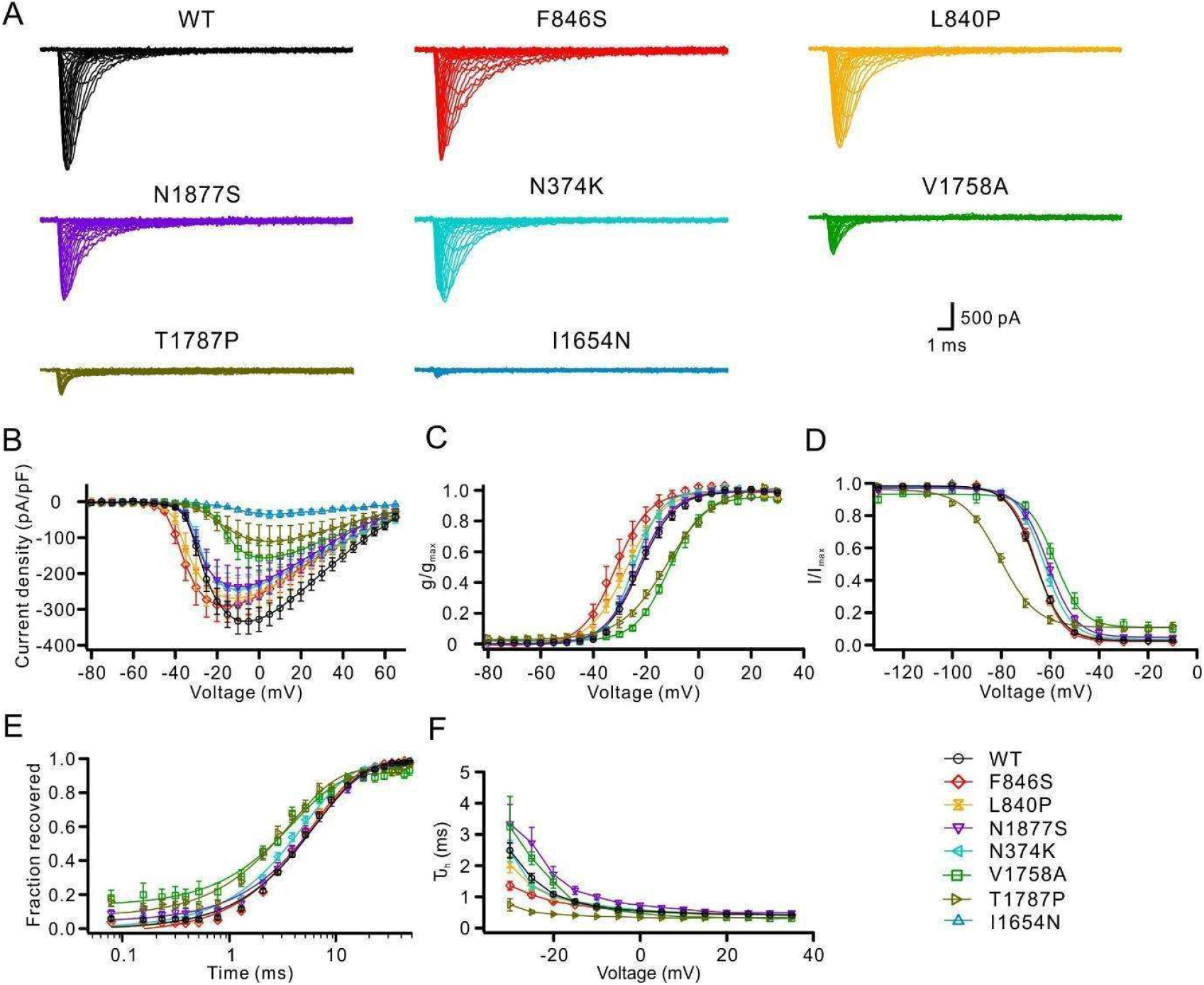
Functional characterizations of *SCN8A* variants in the neuroblastoma cell line ND7/23. WT or mutant Na_V_1.6 channels were transfected into ND7/23 cells and recorded in the presence of TTX to block endogenous Na^+^ channels. (**A**) Representative Na^+^ current traces for transfected Na_V_1.6 wild-type (WT, black) or mutant channels (colour code in the right lower corner). (**B**) Peak Na^+^ currents normalized by cell capacitance were plotted versus voltage. Both the p.(Phe864Ser) and p.(Leu840Pro) variants caused a hyperpolarising shift of the current-voltage relationship, whereas the p.(Val1758Ala) and p.(Thr1787Pro) variants caused a depolarising shift compared to WT channels. P.(Ile1654Asn), p.(Val1758Ala) and p.(Thr1787Pro) variants significantly decreased the current density in comparison to WT. WT: n = 30; mutants: n = 14-19. (**C**) Voltage-dependent steady-state activation curves. Lines represent Boltzmann functions fit to the data points. (**D**) Voltage-dependent steady-state inactivation curves. Lines represent Boltzmann functions fit to the data points. (**E**) Time course of recovery from fast inactivation at -100 mV. The p.(Val1758Ala), p.(Thr1787Pro) and p.(Asn374Lys) variants accelerated the recovery from fast inactivation compared to WT. (**F**) Voltage-dependence of the major time constant of fast inactivation τ_h_. Shown are means ± SEM (**B**-**F**). Numbers of recorded cells and statistical analysis for all experiments are provided in Supplementary Table 2.

**Figure 2.**
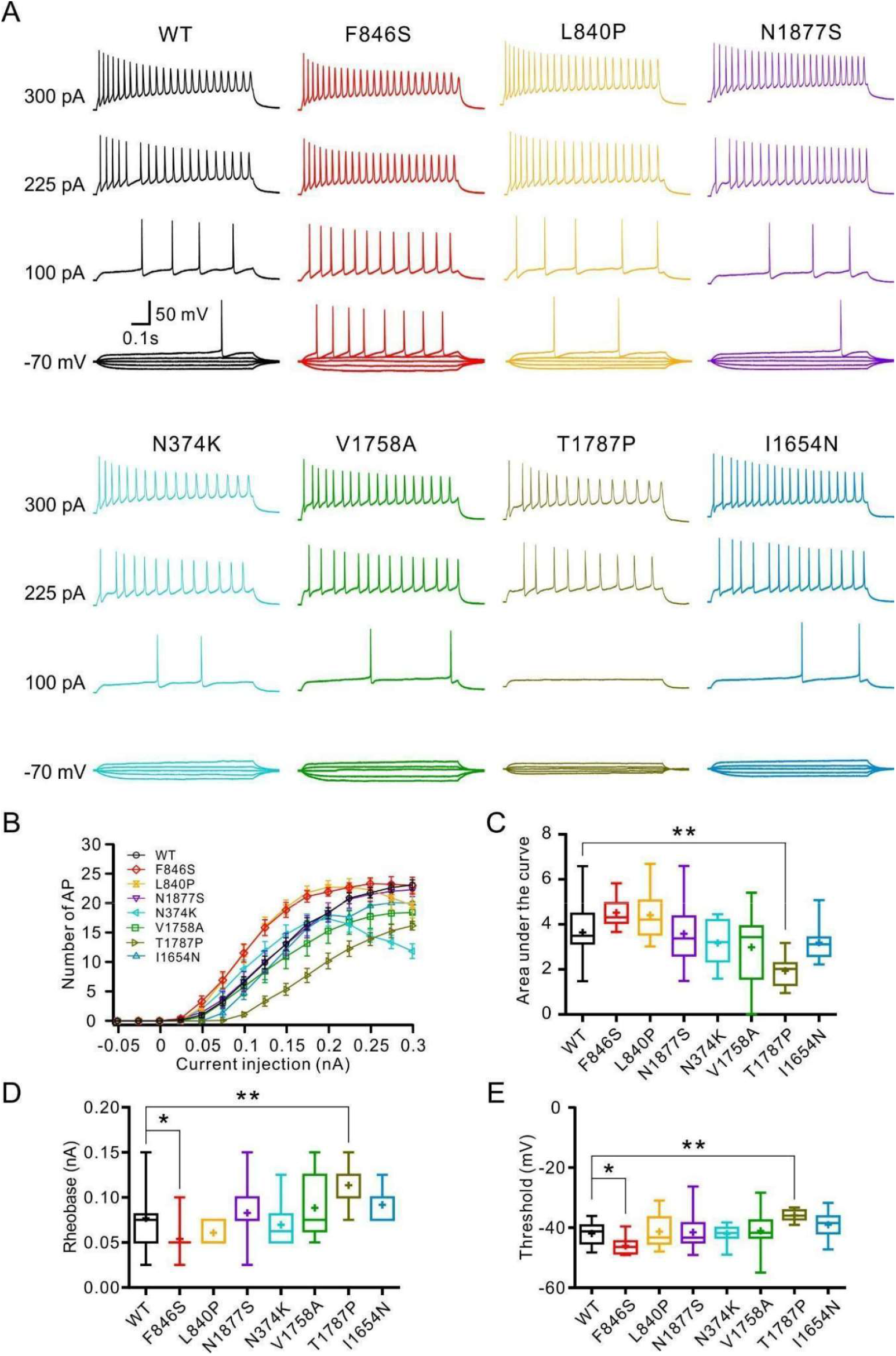
Effects of *SCN8A* variants in primary cultured hippocampal mouse neurons in the absence of TTX. Neurons were transfected with WT or mutant Na_V_1.6 channels and recorded in the absence of TTX. (**A**) Representative voltage traces of evoked action potentials (APs) in the absence of TTX from neurons transfected with WT (black) or mutant neurons (colour code indicated in **B**). (**B**) Numbers of evoked action potentials plotted versus injected current in the absence of TTX. Shown are means ± SEM. (**C**) Area under the curve for the input-output relationships. The p.(Thr1787Pro) variant shows a significantly decreased area under the curve compared to WT channels. (**D** and **E**) Rheobase (**D**) and threshold (**E**) of APs were decreased for neurons transfected with the p.(Phe846Ser) variant, but increased for neurons transfected with the p.(Thr1787Pro) variant compared to WT channels. Box-and-whisker plots (**C**-**E**) show means (plus sign), the 25th, 50th, 75th percentiles, minima and maxima; * p < 0.05; ** p < 0.01; *** p < 0.001; one-way ANOVA with Dunnett′s posthoc test or ANOVA on ranks with Dunn′s posthoc test were performed. Numbers of recorded cells and statistical analysis are provided in Supplementary Table 3.

Next, intrinsic and firing properties were examined in transfected cultured hippocampal mouse neurons in the absence or presence of tetrodotoxin (TTX). In the absence of TTX, action potential (AP) firing was jointly determined by endogenous and transfected Na^+^ channels. Only the p.(Thr1787Pro) variant significantly decreased the AP firing rate compared to neurons transfected with WT channels, as revealed by the area under the curve of input-output relationships (Figs. 2B, 2C, Supplementary Table 3). The p.(Phe846Ser) variant decreased, whereas the p.(Thr1787Pro) variant increased both rheobase and AP threshold compared to the WT (Figs. 2D, 2E, Supplementary Table 3), indicating the former enhanced, whereas the latter impaired neuronal excitability.

In the presence of TTX, AP firing was dependent on the TTX-insensitive transfected Na^+^ channels. Under these conditions, p.(Phe846Ser) and p.(Asn1877Ser) variants significantly increased AP firing, whereas p.(Thr1787Pro) and p.(Ile1654Asn) decreased AP firing (Figs. 3B, 3C, Supplementary Table 4). Additionally, the p.(Phe846Ser), p.(Leu840Pro) and p.(Asn1877Ser) variants decreased rheobase and the former two variants further reduced the threshold for AP firing indicating increased neuronal excitability. In contrast, neurons transfected with p.(Val1758Ala), p.(Thr1787Pro) or p.(Ile1654Asn) variants exhibited significantly decreased peak Na^+^ currents resulting in very few action potentials firing in the presence of TTX, hence no parameters of single APs were obtained. Neurons expressing p.(Asn374Lys) showed comparable Na^+^ peak currents and AP firing to WT channels (Fig. 3, Supplementary Table 4).

**Figure 3.**
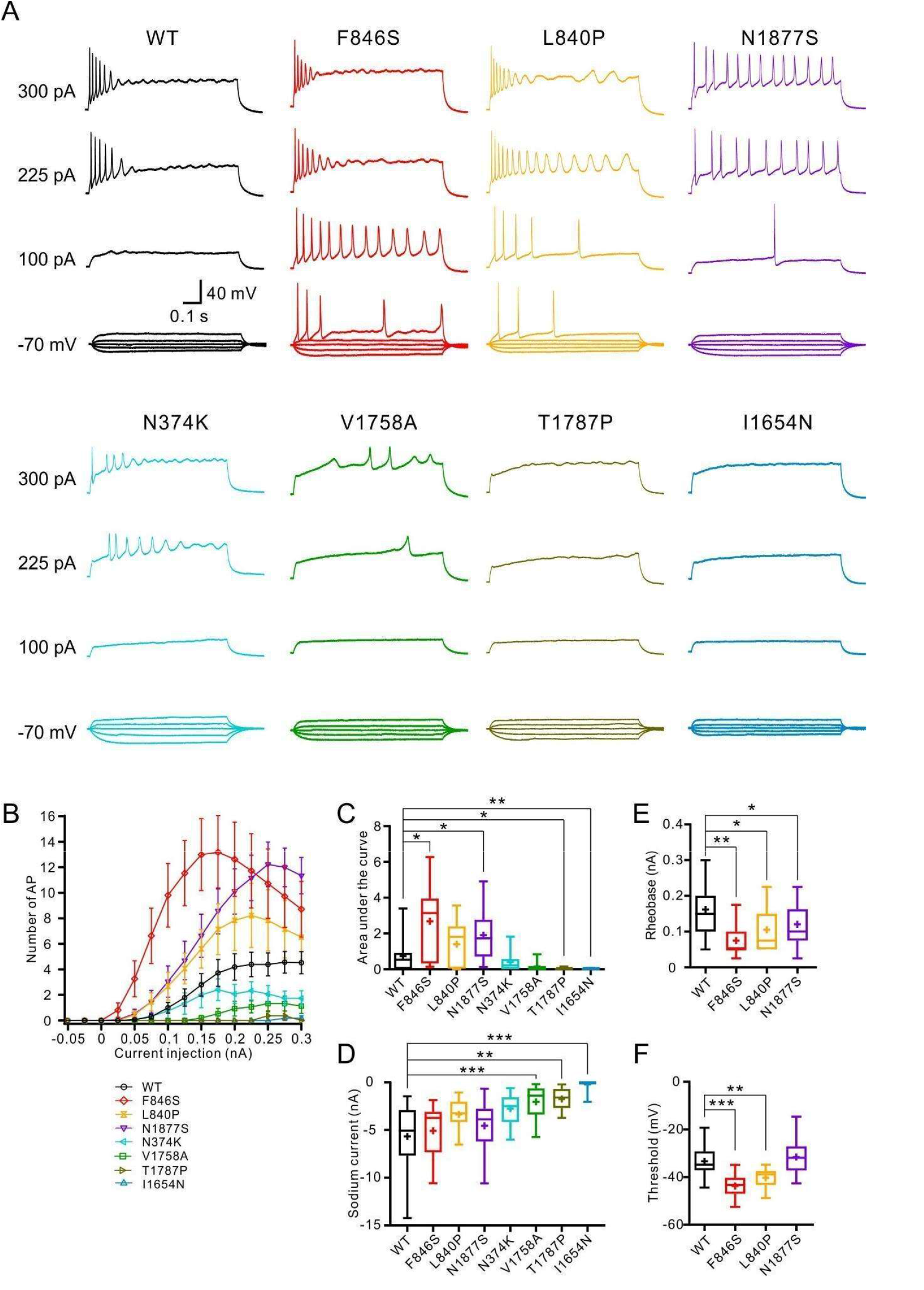
Neuronal properties carried only by transfected WT or mutant Na_V_1.6 channels. Hippocampal neurons transfected with wild-type or mutant Na_V_1.6 channels were recorded in the presence of TTX to block endogenous Na^+^ channels. (**A**) Representative voltage traces of evoked action potentials (APs) from neurons transfected with WT (black) or mutant channels (colour code in the lower left corner). (**B**) Numbers of evoked action potentials plotted versus injected current in the presence of TTX. Shown are means ± SEM. (**C**) Area under the curve for the input-output relationships. (**D**) Peak Na^+^ current amplitudes of neurons transfected with WT or mutant Na_V_1.6 channels in the presence of TTX. (**E** and **F**) Rheobase (**E**) and Threshold (**F**) were significantly decreased in neurons transfected with p.(Phe846Ser) or p.(Leu840Pro) variants compared to WT channels. The p.(Asn1877Ser) variant also significantly decreased the rheobase. The rheobase or threshold could not be obtained in neurons transfected with p.(Asn374Lys), p.(Val1758Ala), p.(Thr1787Pro) and p.(Ile 1654Asn) mutant channels due to very few evoked APs. Box-and-whisker plots (**C**-**F**) show means (plus sign), the 25th, 50th, 75th percentiles, minima and maxima; * p < 0.05; ** p < 0.01; *** p < 0.001; one-way ANOVA with Dunnett′s posthoc test or ANOVA on ranks with Dunn′s posthoc test were performed. Numbers of recorded cells and statistical analysis are provided in Supplementary Table 4.

In summary, p.(Phe846Ser), p.(Leu840Pro) and p.(Asn1877Ser) showed a GOF effect or increased neuronal firing; p.(Asn374Lys) showed mild GOF effects and did not alter neuronal firing; p.(Val1758Ala), p.(Thr1787Pro) and p.(Ile1654Asn) showed LOF effects or decreased neuronal firing.

### Genotype-phenotype correlations in GOF and LOF variant carriers and extrapolation to the whole cohort of affected individuals

Clinical data combined with functional results of this and earlier studies enabled us to explore genotype-phenotype correlations. Since there are 165 affected individuals carrying a known LOF (n=32) or GOF (n=133) variant, a detailed phenotypic analysis in those affected individuals and a comparison to the whole cohort may allow an extrapolation of genotype-phenotype correlations that could be valid for all individuals in which causative *SCN8A* variants have been detected.

### Phenotypes of affected individuals with loss-of-function variants

Thirty-two affected individuals (12 previously unpublished) carried clear LOF variants, either deletions, in-frame deletions, splice-site, frameshift or stop-variants, or missense variants with confirmed LOF effect either in this or previous studies^17-19, 21^. Twenty-one affected individuals had epilepsy. The median age at seizure onset was 28.5 months (range 1 month to 14 years). Seizure types included absence seizures (11/21), GTCs (7/21) and myoclonic seizures (5/21). Additionally, 5/21 had febrile seizures within the first year of life, all with later onset (>two years of age) of additional seizure types. Fourteen/32 had a phenotype of generalized epilepsy, 11/32 had a neurodevelopmental disorder without epilepsy, two/32 had DEE and the remaining five/32 had an unclassifiable epilepsy. EEGs showed background slowing, generalized spike-wave activity, slow rhythmic activity or remained normal. Nine/21 affected individuals with epilepsy were seizure-free: one with an SCB (LTG), seven with non-SCBs (VPA monotherapy in four affected individuals) and one on a combination (LTG and TPM). Out of four additional affected individuals who tried SCBs, one had a worsening of seizures with ZNS, two had no effect, and one had a reduction of seizures with LTG.

None of the affected individuals with LOF variants died prematurely.

When we compared this presentation of clear LOF variant carriers to the phenotypes of the whole cohort of affected individuals, there was a clear correlation to two groups; one with generalized epilepsy, and one with neurodevelopmental disorder without epilepsy. The only exception was the two affected individuals with DEE in the LOF group. The main phenotypic features of these groups including an analysis of the age of onset of seizures (except febrile seizures) are summarized and contrasted in Figure 4 and Table 1.

**Figure 4.**
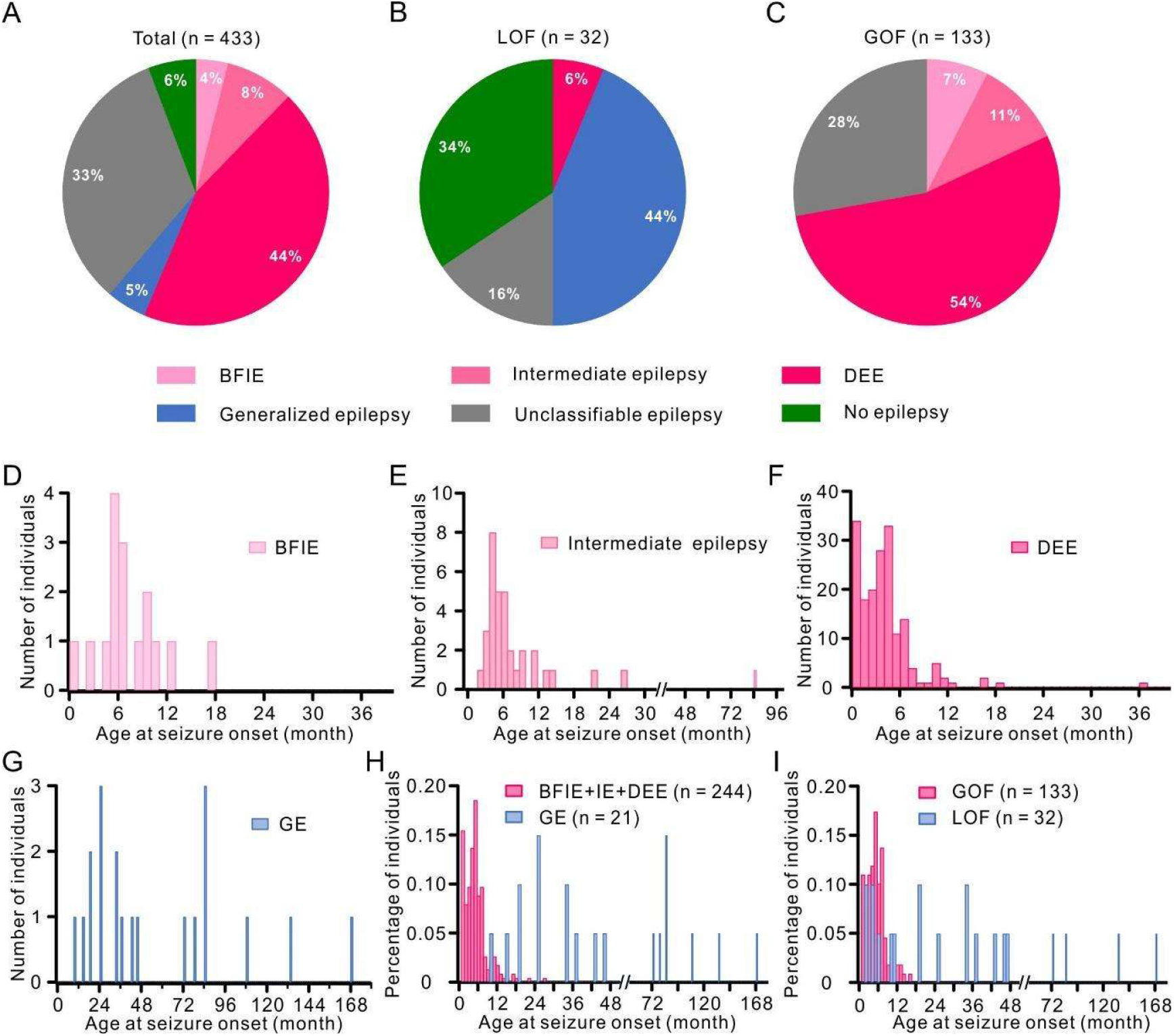
Distribution of affected individuals carrying *SCN8A* variants according to phenotype and age of seizure onset. (**A**-**C**) Phenotypic subgroups of the total cohort (**A**), and individuals carrying LOF (**B**) or GOF (**C**) variants. (**A**) In the total cohort, 244 affected individuals (56.4%) had BFIE, IE or DEE; 21 individuals (4.8%) had GE; 25 individuals (5.8%) had no epilepsy. (**B**) Twenty-five affected individuals had GE or no epilepsy, accounting for 78.1% of LOF variant carriers, (**C**) whereas 72.2% of GOF variant carriers had BFIE, IE or DEE. (**D**-**I**) Histogram of the age at seizure onset in affected individuals with BFIE(**D**), IE (**E**), DEE (**F**), GE (**G**), BFIE+IE+DEE versus GE (**H**) and GOF versus LOF variant carriers (**I**). Affected individuals with BNIE, IE or DEE exhibited an earlier median age of seizure onset than individuals with GE (**D**-**H**), which is also observed for GOF vs. LOF variant carriers (**I**). Histogram bin size = 1 month. BFIE=Benign Familial Infantile Epilepsy; IE=Intermediate Epilepsy; DEE= Developmental and Epileptic Encephalopathy; GE= Generalized Epilepsy; GOF= Gain-of-function; LOF=Loss-of-function.

### Phenotypes of affected individuals with gain-of-function variants

One hundred and thirty-three affected individuals from the total cohort (30 unpublished) carried missense variants that were shown to cause GOF^1, 6, 17, 19, 29, 30^. All 133 suffered from epilepsy. The most prevalent seizure types were focal (31.6%), focal tonic (31.6%) or bilateral TC seizures (42.9%). Febrile seizures were seen in 4.5%. Median age at seizure onset was four months (range: first day of life to 45 months). Half of the affected individuals (54.1%) were diagnosed with DEE, followed by unclassifiable epilepsy in 27.8%, intermediate epilepsy in 10.5% and BFIE in 7.5%. Individuals with BFIE had either no interictal epileptiform abnormalities or rare diffuse spike and wave complexes. Individuals with intermediate epilepsy had heterogeneous EEG features with trains of beta and delta activity, and focal spike and slow waves, bilaterally in the parieto-occipital regions, with or without diffuse spreading. The majority of individuals with severe DEE had background slowing, polymorphic delta and beta activity, and multifocal spike and slow waves, predominant in the posterior quadrants.

Treatment data was available in 84 affected individuals. Twenty-six/84 affected individuals were seizure free; 16 with SCBs. Seizure reduction was seen in 47 affected individuals, 18 with SCBs. Two affected individuals showed worsening of seizures with SCBs; both affected individuals were resistant to several ASMs and had increased seizure frequency with OXC and LEV^31^. Eight affected individuals did not try SCB treatment: two had a mild phenotype and were seizure free with VPA or VPA+LEV^5, 32^ and one affected individual also had a mild phenotype and seizure reduction with VPA^33^. The remaining five affected individuals all had pharmacoresistant seizures, it is unknown why SCBs were not tried.

Twelve affected individuals were prematurely deceased (9.0%), all with a DEE phenotype; ten due to a general worsening in their overall and neurological condition followed by organ failure and two due to definite or probable SUDEP (1.5%).

As in the LOF group, there was a clear correlation in the GOF group to specific phenotypes in the whole cohort, namely BFIE, intermediate epilepsy, and DEE. None of the GOF variant carriers had a generalized epilepsy or a neurodevelopmental disorder without epilepsy.

A summary of these data is provided in Figure 4 and Table 1. The obvious difference in the age of onset was statistically significant between GOF and LOF variant carriers, and between BFIE/IE/DEE vs. GE in general. Statistical analysis also confirmed that affected individuals with LOF variants mostly had a generalized epilepsy, whereas affected individuals with GOF had one of the focal epilepsy phenotypes (BFIE, intermediate epilepsy or DEE).

### Correlation of clinical and electrophysiological phenotypes in the GOF group

We scored all GOF variants according to their functional characterizations from this and previous studies, including a total of 16 variants. The LOF variants were not included due to a large proportion of deleterious variants, which lead to non-functional protein that cannot be well differentiated. The electrophysiological score was determined by the effect of biophysical changes of variants on neuronal firing simulated by a single-compartment conductance based model (see methods and supplementary methods). This analysis revealed that (i) alterations of activation properties severely affect neuronal firing; (ii) changes in the sodium conductivity have a moderate effect; (iii) changes of inactivation properties and the persistent current affect neuronal firing mildly (Fig. 5A-F, Supplementary Table 5).

**Figure 5.**
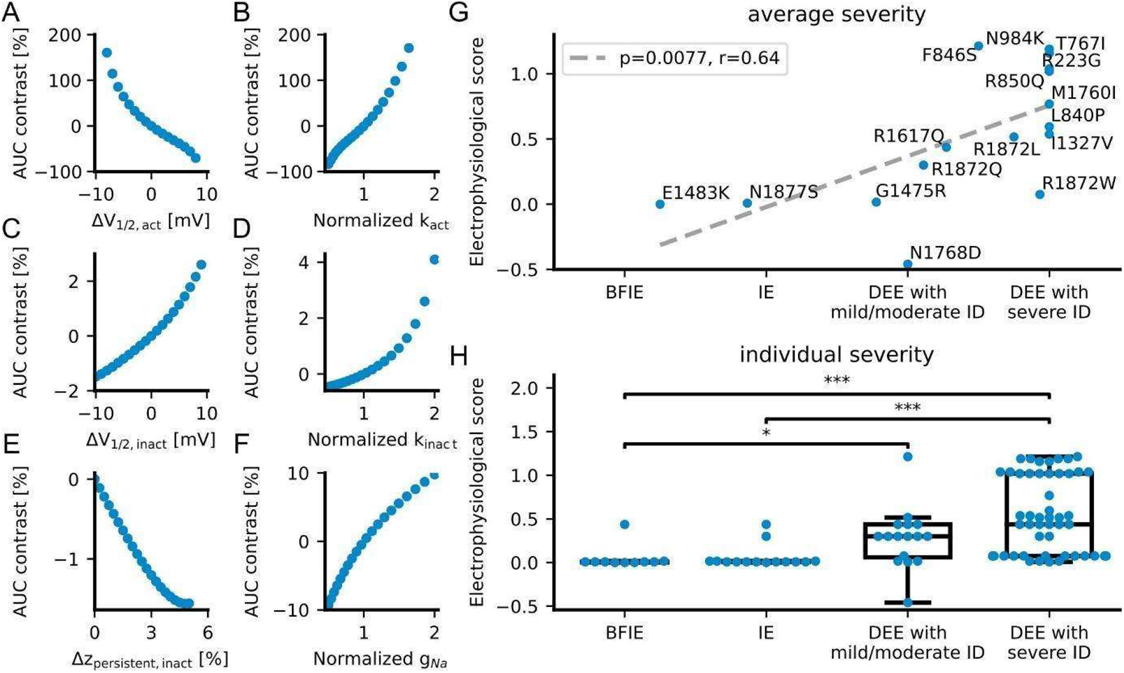
Correlation of a computed electrophysiological score with the clinical severity of *SCN8A* GOF variants. Electrophysiological scores of *SCN8A* GOF variants were obtained according to the effect of variants on AP firing simulated by a single-compartment conductance based model. (**A**-**F**) The contrast of the simulated area under the input-output curve (AUC, equation 1) as a function of the changes of single Na^+^ current gating parameters, such as: (**A**) the V_1/2_ of the activation curve; (**B**) the slope factor k of the activation curve; (**C**) the V_1/2_ of the fast inactivation curve; (**D**) the slope factor k of the fast inactivation curve; (**E**) the persistent Na^+^ current; (**F**) the Na^+^ conductivity (or current density). Changes of the V_1/2_ and the slope of the activation curve had a much stronger effect on the AUC than other parameters. (**G**) Correlation (dashed gray line) of the simulation-based score (equation 9) with the severity of each *SCN8A* GOF variant averaged over affected individuals (blue dots with respective one-amino acid code). (**H**) Distributions of simulation-based scores of each patient (blue dots) for each of the four categories of clinical severities. * p < 0.05; *** p < 0.001 (ANOVA on ranks with Dunn′s posthoc test).

The electrophysiological scores of GOF variants clearly correlated with the clinical severity of affected individuals carrying these variants (Fig. 5G, p = 0.0077, r = 0.64), which were previously classified into BFIE, IE, DEE with mild/moderate ID and DEE with severe ID (scored as 1, 2, 3 and 4 respectively). Variants causing different phenotypes were averaged over this score (see methods). In a separate analysis, we also compared the individual severity of each affected individual with the electrophysiological score of the variants. The scores in the groups with DEE are significantly higher than the ones in BFIE and IE, but there is no significant difference between BFIE and IE or DEE with mild/moderate ID and DEE with severe ID (Fig. 5H).

### Genetic landscape of *SCN8A* variants

We analyzed 297 variants in all 433 individuals. Missense variants (n=273) accounted for the majority of disease-causing variants (91.9%). Twenty-one variants were deleterious, either frameshift, stop or splice-site variants. Three patients had biallelic missense variants^34^. Sixty-one recurring missense variants were found in 244 affected individuals. Two hundred and twenty-three variants were *de novo*, 30 were inherited from an affected or unaffected mosaic parent, and for 180 segregation was unknown.

The distribution of variants across the Na_V_1.6 channel protein is shown in Figure 6. It is remarkable that most GOF variants (shown in red) and also variants causing focal epilepsy (BFIE/IE/DEE) without functional data (shown in light red), are concentrated at the cytoplasmic side and in the voltage sensors, whereas many LOF variants are found in the pore region. This is also illustrated in a 3D structural model which also reveals that common variants taken from the gnomAD database are located in different regions. Most disease-causing variants were located in functionally important and evolutionary conserved regions of the channel, whereas variants from the gnomAD database are in less conserved regions (Supplementary. Fig. 1 and Supplementary video 1).

**Figure 6.**
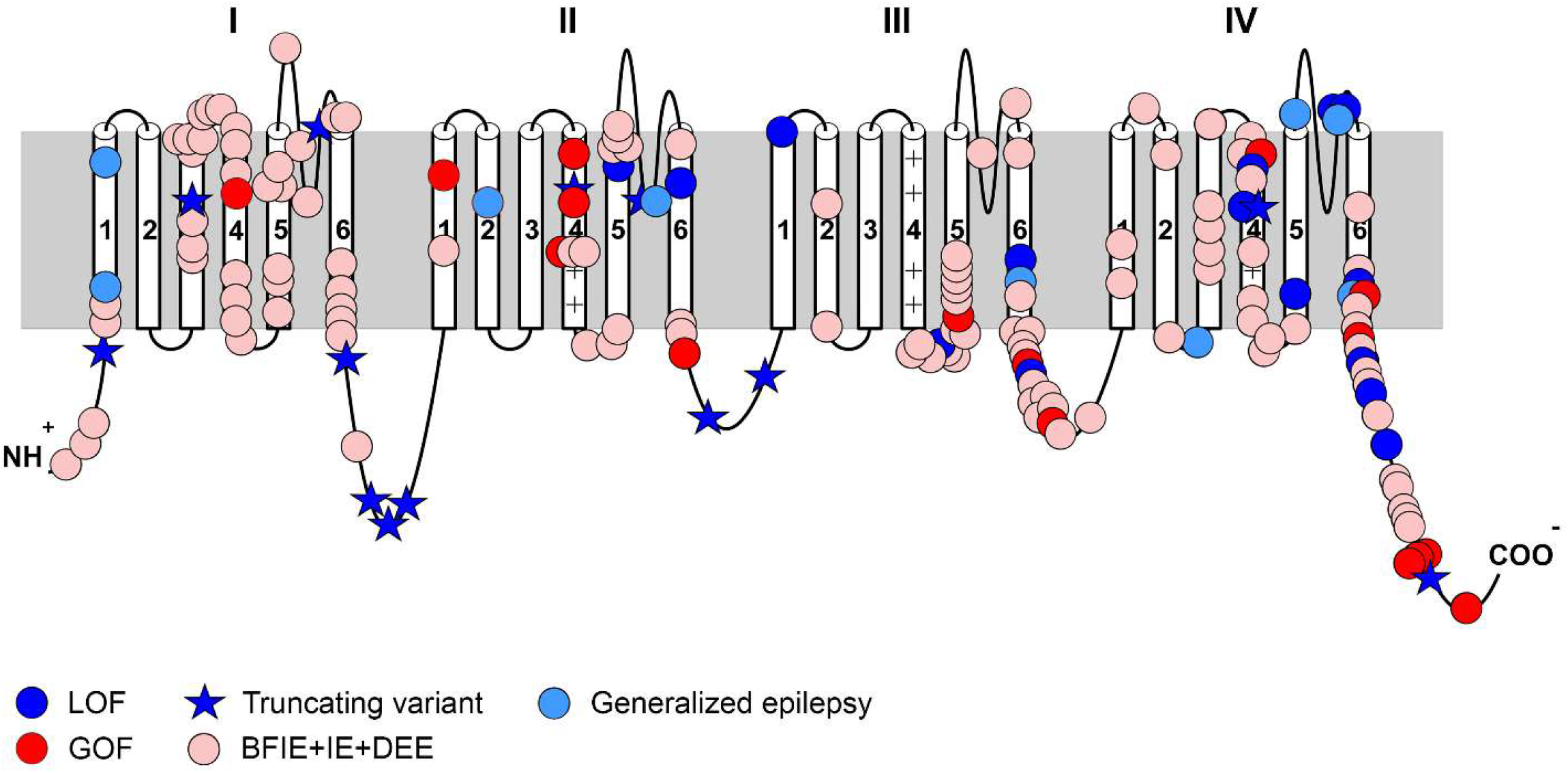
Location of *SCN8A* variants associated with neurodevelopmental disorders. Schematic 2D representation of the *SCN8A* gene displaying the location of pathogenic variants, showing clustering of GOF variants and BFIE+IE+DEE phenotypes in the transmembrane regions, especially the voltage-sensor, while LOF variants and GE phenotypes are mainly seen in the pore regions. Abbreviations: BFIE=Benign Familial Infantile Epilepsy; IE=Intermediate Epilepsy; DEE= Developmental and Epileptic Encephalopathy; GE= Generalized Epilepsy; GOF= Gain-of-function; LOF=Loss-of-function.

### Frequency of *SCN8A*-related disorders in the Danish population

From the year 2006 to 2017, an average of 60,934 children were born per year. During the same period, 13 affected individuals were diagnosed with an *SCN8A*-related disorder in Denmark, yielding an estimated frequency of 1/56,247.

### Evaluation of treatment effects

Treatment responses to specific ASMs were grouped into four categories - seizure-free, seizure reduction, no change and worsening - as described in the methods section. We differentiated between SCBs and non-SCBs, in LOF and GOF variant carriers, GE and focal epilepsies (BFIE/IE/DEE). The results are shown in Figure 7. GOF variant carriers responded better to SCBs than to non-SCBs (Fisher’s Exact Test, p < 0.001), whereas there was no difference between the treatment of SCBs and non-SCBs for LOF variant carriers (numbers very small, Fisher’s Exact Test, p = 0.48). Similar results were obtained for the treatment of SCBs versus non-SCBs in affected individuals with focal epilepsy vs. those with GE.

**Figure 7.**
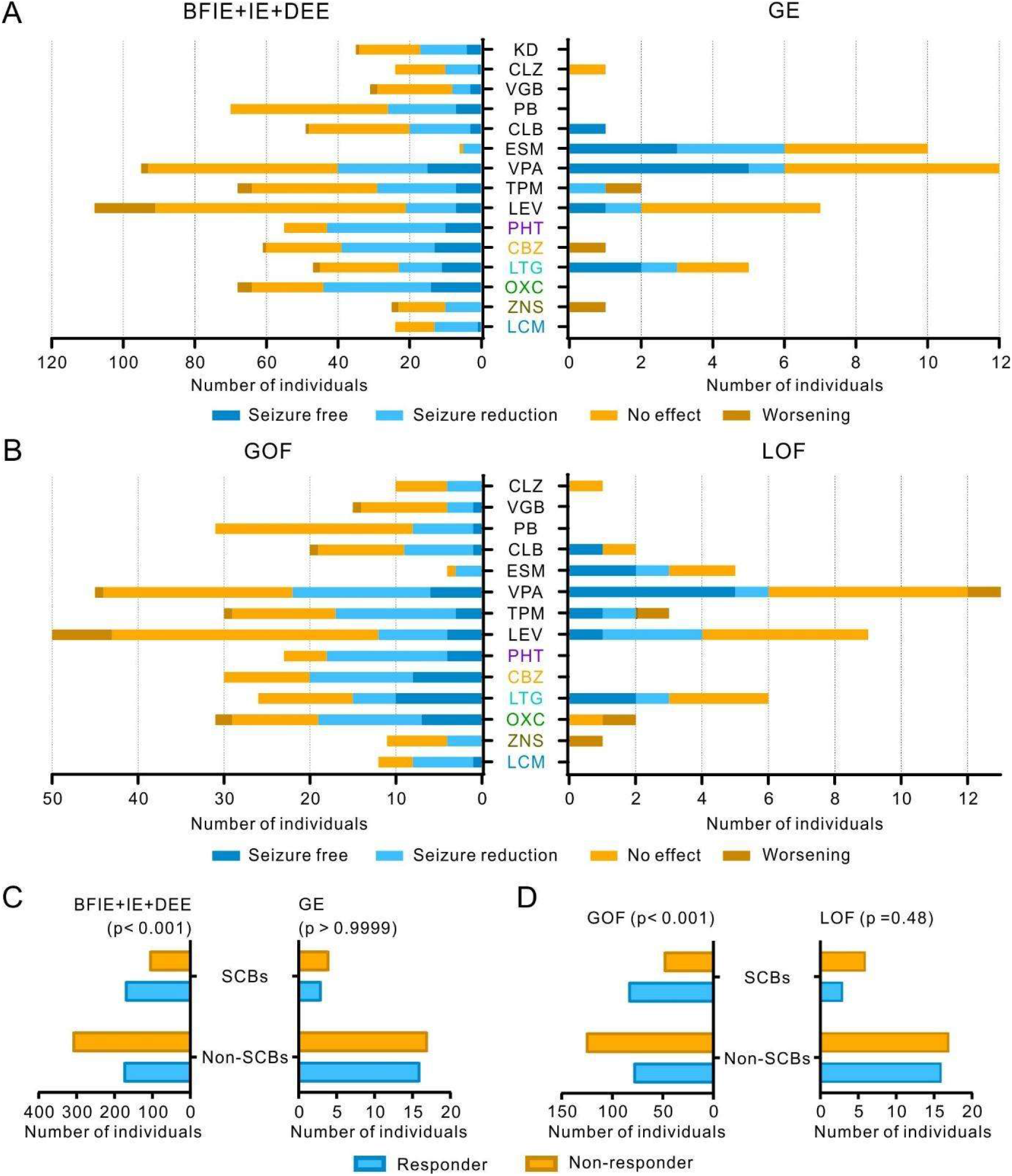
Treatment responses to anti-seizure medications in affected individuals carrying *SCN8A* variants. (**A, B**) Treatment effects of anti-seizure medications (ASM) on seizures in affected individuals with BFIE, IE or DEE versus those with GE (**A**) and those carrying *SCN8A* GOF vs. LOF variants (**B**). KD = ketogenic diet; CLZ = clonazepam; VGB = vigabatrin; PB = phenobarbital; CLB = clobazam; ESM = ethosuximide; VPA = valproate; TPM = topiramate; LEV = levetiracetam; PHT = phenytoin; CBZ = carbamazepine; LTG = lamotrigine; OXC= oxcarbazepine; ZNS = zonisamide; LCM = lacosamide. PHT, CBZ, LTG, OXC, ZNS and LCM are sodium channel blockers (SCBs). (**C, D**) Individuals with BFIE, IE or DEE and those carrying *SCN8A* GOF variants responded significantly better to SCBs than non-SCBs, whereas treatment of SCBs or non-SCBs did not cause a different effect in individuals with GE and those carrying *SCN8A* LOF variants (please consider the very small numbers in these latter categories). P values derived from Fisher’s exact test are provided in the Figure. Responders were defined as those becoming seizure free or experienced a seizure reduction staying on the drug; non-responders were defined as those experiencing no effect or seizure worsening.

## Discussion

Our study provides a detailed analysis of the correlation between clinical phenotypes, genotypes and electrophysiological characterizations, suggesting the following four main conclusions:

1. There are five main phenotypes in *SCN8A*-related disorders in this largest series of 433 individuals collected to date: BFIE, IE, DEE, GE and neurodevelopmental disorder without epilepsy.
2. There is a clear correlation between GOF variants causing focal epilepsy of different/increasing severity (BFIE, IE, DEE) and LOF variants causing GE or neurodevelopmental disorders without epilepsy, allowing an extrapolation from the clinical phenotypes of clear GOF and LOF variant carriers to the whole cohort. There were only two exceptions from this rule: two DEE affected individuals with early onset carrying truncating LOF variants (see discussion below for exact phenotypes).
3. The clinical severity of epilepsies associated with GOF variants is at least partially determined by the degree of the electrophysiological dysfunction, as there was a significant correlation between the clinical and electrophysiological phenotypes using a new system to estimate the influence of different gating parameters on neuronal firing.
4. As previously suggested in smaller studies, seizures in individuals with disease-causing GOF variants respond better to sodium channel blockers (SCBs) than to other ASMs.

### Functional studies

We examined altogether 14 variants (this study and ^19^ covering the whole clinical spectrum of *SCN8A*-related phenotypes. Using both neuroblastoma cells to study the effects of the mutant channels on gating properties and murine primary neuronal cultures to study effects on intrinsic neuronal properties and firing, we found mild GOF effects or an increase in neuronal firing to be associated with BFIE or IE (p.(Gly1475Arg), p.(Glu1483Lys) and p.(Asn1877Ser)), whereas more severe gating defects, particularly strong hyperpolarizing shifts of steady-state activation that also on average led to stronger increase in neuronal firing, were associated with DEE (p.(Arg223Gly), p.(Leu840Pro), p.(Phe846Ser), p.(Met1760Ile) and p.(Arg1872Trp)). One previously published variant, p.(Arg223Gly), was initially suggested to cause a predominant LOF^11^ and had a phenotype of early onset spasms, severe ID and an epileptic encephalopathy. However, there were also GOF features identified, and when we retested this variant in our system, GOF effects predominated and it increased neuronal firing (Liu, Koko, Lerche, unpublished). When we used a one compartment neuronal model system, we were able to weight the gating defects of all GOF variants that were functionally investigated so far according to their effect on neuronal firing, revealing a significant correlation between the severity of the clinical and electrophysiological phenotypes. Such a correlation was not observed when we used a different scoring system (data not shown) based only on the degree of the electrophysiological dysfunction (i.e. without weighting the effects on neuronal firing), which, however, served well previously to correlate clinical and electrophysiological phenotypic severity in *SCN2A*-related epilepsy^35^.

In contrast, the p.(Ile1654Asn), p.(Thr1787Pro) and p.(Val1758Ala) variants, all causing GE with absence seizures as a new recurring phenotype, showed a LOF in neuroblastoma cells or decreased neuronal firing. Also, variants not causing epilepsy either caused a clear LOF and decreased neuronal firing (p.(Gly964Arg) and p.(Arg1620Leu)), or a strong GOF on channel gating leading to a depolarisation block, i.e. LOF on a neuronal level (p.(Ala1622Asp))^19^. Additional variants characterized by other groups confirm these results. Many other variants causing GOF effects mainly on channel gating (only few studies with limited results on neuronal firing) were clearly associated with focal epilepsies (Supplementary Table 1).

The p.(Ala1319Thr) variant caused a depolarising shift of the activation curve of Na^+^ current in *x. laevis* oocytes^36, 37^ and decreased firing of cerebellar Purkinje cells^38, 39^ indicating a LOF. The *Scn8a Med*^*jo*^ mouse carrying the corresponding variant in the mouse channel exhibited absence epilepsy revealed by slow-wave discharges (SWD) in EEG recordings associated with absence-like seizures^40^, reproducing the clinical phenotype found in the affected individual carrying the p.(Ala1319Thr) variant. Other epilepsy-related LOF variants were also associated with absence epilepsy, such as p.(Asn544fs*39) and p.(Glu587*)^5, 31^. In another, conditional mouse model, a complete *Scn8a* LOF restricted to inhibitory neurons of the thalamic reticular nucleus also caused absence-like epilepsy with generalized SWD^40, 41^. It is therefore intriguing to speculate that *SCN8A* variants cause absences and other generalized seizures due to a LOF in inhibitory neurons, whereas the GOF effects are more important in excitatory neurons, as discussed in more detail further below including the clinical consequences.

The p.(Asn374Lys) variant showed only very mild GOF gating defects but did not change parameters in neurons. The phenotype with late onset focal epilepsy at seven years without other neuropsychiatric symptoms does also not fit to the five main categories. Thus, while we cannot exclude a mild contribution of this variant to seizures in this affected individual, we rather consider it to be a benign polymorphism.

### Phenotype correlations in GOF vs. LOF variant carriers

When we compared the clinical phenotypes of the confirmed 133 GOF and 32 LOF variant carriers with the whole group of 433 affected individuals, there were strikingly congruent phenotypes, which strongly suggest that BFIE, IE and DEE are generally caused by GOF, while GE and neurodevelopmental disorders without epilepsy are caused by LOF variants. Not only did seizure types resemble each other in those groups, but also an analysis of the age of onset of seizures strongly confirmed this hypothesis (Table 1, Fig. 4). Only two cases with presumed DEE and LOF variants did not fit into this pattern: one (p.(Pro1428_Lys1473del)) with onset of pharmacoresistant hemiclonic and tonic seizures at ten months of age, severe ID, loss of eye contact, severe extrapyramidal movement disorder, gastroparesis and microcephaly^14^, and another affected individual (p.(Met1481Ilefs*12)) with onset of pharmacoresistant tonic, GTC seizures and episodes of status epilepticus at five months of age, severe ID, no speech and poor eye contact. EEG showed multifocal epileptic discharges in both cases. Thus, those two cases could not be differentiated from the DEE cohort caused by GOF variants.

In a previous study on *SCN2A*^*42*^, we demonstrated that different early-onset epilepsy syndromes were caused by GOF variants, whereas later onset seizures were due to a LOF of Na_V_1.2. In *SCN2A*, the boundary for the onset of seizures between GOF and LOF was very early – approximately three months of age^42^. In the present study, the onset in both groups was later (median age at onset of four months in the GOF subgroup, and 28.5 months in the LOF subgroup), and the ranges were more overlapping. This difference may be a reflection of the differential developmental expression of *SCN2A* vs. *SCN8A*, as *SCN2A* is expressed neonatally and earlier than *SCN8A*^*43, 44*^. An additional difference is that *SCN2A* variant carriers have better defined electroclinical epilepsy syndromes. Both groups share a phenotype of BFNIE/BFIE, with a later onset in *SCN8A*-related epilepsy. Some individuals carrying *SCN8A* variants develop paroxysmal kinesigenic dyskinesia^3^, whereas *SCN2A* variant carriers may develop a form of later onset episodic ataxia^45^. There were also differences regarding the syndromes of LOF variant carriers in both genes. Generalized epilepsy was not generally observed in *SCN2A*, although absence epilepsy has been reported in mice with complete LOF of *Scn2a*, when knocked out in excitatory neurons^46^. Instead, West syndrome was a common phenotype in human *SCN2A*, but also phenotypes with generalized seizures and EEG abnormalities were found in the LOF group (epilepsy with myoclonic-atonic seizures or Lennox-Gastaut syndrome). For *SCN8A*, the phenotypes were rather unspecific generalized epilepsy, mainly with absence seizures. However, we found that a LOF subgroup had a phenotype resembling *SCN1A* LOF, with febrile seizures preceding additional seizure types, resembling a generalized/genetic epilepsy with febrile seizure plus (GEFS+) phenotype. *SCN8A* is expressed in both excitatory and inhibitory neurons^47^, which may well explain the phenotypic similarity between some of the *SCN1A*- and *SCN8A*-related phenotypes, since *SCN1A* is considered to be the main sodium channel in inhibitory GABAergic neurons^48, 49^. Additionally, it has been found that knockout of *Scn8a* in mice decreased cortical excitability resulting in convulsive seizure protection^40^. Furthermore, mice with global reduction of *Scn8a* had fewer seizures compared to mice with *Scn8a* deletion only in inhibitory neurons^41^. In contrast, mice carrying a GOF *Scn8a* variant (p. (Arg1872Trp)) exhibited convulsive seizures and premature death; however, activation of this variant only in inhibitory neurons did not induce seizures or overt neurological dysfunctions^20^. Altogether, *SCN8A* GOF variants may mainly cause hyperexcitation in excitatory neurons leading to TC and focal seizures, whereas *SCN8A* LOF variants mainly induce hypoexcitation in inhibitory neurons causing generalized and particularly absence seizures. Further studies may elucidate the distinct roles of *SCN8A* and the other sodium channel genes in different neuronal circuits.

### Genetic landscape

The vast majority of the detected variants were missense, and we found several recurrent variants. Those detected in more than ten affected individuals include p.(Arg850Gln/Gly) seen in 12 affected individuals, p.(Gly1475Arg) seen in 19 affected individuals, p.(Arg1617Trp/Gln/Leu) seen in 18 affected individuals, p.(Arg1872Trp/Gln/Leu) seen in 38 affected individuals and p.(Asn1877Ser) seen in 23 affected individuals. All of these variants cause a clear GOF. Why some of these variants, especially p.(Gly1475Arg) and p.(Arg1617Trp/Gln/Leu), have a high phenotypic variability and others not, remains to be elucidated. Other genetic or environmental factors might play an important role. Additionally, we found families with intra-familial variability, such as the family of proband #85, whose mother and two older siblings have BFIE and are seizure-free with normal intellect, whereas the proband suffers from pharmacoresistant DEE.

### Estimated frequency

We found the frequency of *SCN8A* related disorders in the Danish population to be 1/56,247, which is higher than for *SCN2A* (1/78,608)^42^ but lower than for *SCN1A* (1/22,000)^50^. Since these three genes are of similar size and homology, we would assume a similar variant frequency. Large-scale genetic testing in isolated ASD cohorts have failed to detect large numbers of *SCN8A* affected individuals^51^, suggesting that a large number of undiagnosed *SCN8A*-ASD affected individuals does not exist and that this is not the cause behind the discrepancy in numbers. It could be that some individuals with truncating/LOF variants are too mildly affected (learning disabilities, mild ID etc.) to be candidates for genetic testing, but are also not healthy enough to be part of control populations (such as blood donors), as reflected in the almost complete absence of truncating variants in gnomAD (pLi score:1). Intra-uterine death in *SCN8A-*related disease could be a third cause not investigated so far.

### Treatment implications

In general, and as expected, affected individuals with GOF effects showed an overall positive response to treatment with SCBs, as has been hypothesized previously^22^. However, many of the affected individuals with DEE still have pharmacoresistant epilepsy (80.4%), underlining the severity of *SCN8A*-related epilepsy. A recent study evaluated the effect of a novel SCB: GS967, which primarily targets the elevated persistent currents without affecting peak currents, as well as having increased potency compared to PHT^52^. Long-term treatment with GS967 was shown to protect *Scn8a*^N1768/+^ mice against premature death, and also alleviated seizure burden^52^. GS967/PRAX-330 (https://adisinsight.springer.com/drugs/800050600) and another new drug - XEN901 (https://clinicaltrials.gov/ct2/show/NCT03467100), specifically targeting *SCN8A* with a goal of rectifying the effects of GOF variants, are currently in Phase I clinical trials. Furthermore, anti-sense oligonucleotide therapy was recently shown to prolong survival in *Scn8a*^R1872W/+^ (GOF) mice^53^. These novel compounds provide promising advances for precision medicine in *SCN8A* affected individuals. As functional testing is not yet readily available for all affected individuals, it will be important to have clinical markers that may predict the underlying functional effect, when specific blockers of the Na_V_1.6 channel become available on the market. Our study provides an important contribution in this direction, as genotype-phenotype correlations revealed a clearly differential pattern for both GOF and LOF variants with very few exceptions.

## Data Availability

Data will be made available upon request

## Acknowledgements

We thank the affected individuals and their families for participating in this study. The authors wish to acknowledge the resources of MSSNG (www.mss.ng), Autism Speaks and The Centre for Applied Genomics at The Hospital for Sick Children, Toronto, Canada. We thank the Epi25 Collaborative for providing the SCN8A variants in three individuals (supporting grants see below). We also thank the participating families for their time and contributions to this database, as well as the generosity of the donors who supported this program.

## Funding

This work was supported by the German research foundation and the Fonds Nationale de la Recherche in Luxembourg (Research Unit FOR-2715, DFG grants Le1030/15-1 and /16-1 to HL, He8155/1-1 to UBSH, He5415/7-1 to IH, FNR grant INTER/DFG/17/11583046 to PM), the German Federal Ministry for Education and Research (BMBF, Treat-ION, 01GM1907A to HL, DL and PM), the foundation no epilep (to HL to partially support LS), the Medical Faculty of the University of Tuebingen (the Fortüne program 2430-00 to SL). KS, PL, MV were funded by grant: MH CR AZV NU20-04-00279. Epi25 was supported by the National Human Genome Research Institute (NHGRI) grants UM1 HG008895 and 5U01HG009088-02, and the Stanley Center for Psychiatric Research at the Broad Institute. SI was employed by and received a salary from Ambry Genetics. SS hold the GlaxoSmithKline Endowed Chair in Genome Sciences at the Hospital for Sick Children and University of Toronto. P.S. has received speaker fees and participated at advisory boards for Biomarin, Zogenyx, GW Pharmaceuticals, and has received research funding by ENECTA BV, GW Pharmaceuticals, Kolfarma Srl., Eisai. EM was funded by grant: NIH NINDS K08 NS097633 and Foerderer Award for Excellence from The Children’s Hospital of Philadelphia Research Institute. PV received a speaker fee from Nutricia GmbH, Dr. Schär AG / SPA, Eisai. Nestle’, Pediatrica. FZ and PS developed this work within the framework of the DINOGMI Department of Excellence of MIUR 2018-2022 (legge 232 del 2016). IH was also supported by The Hartwell Foundation through an Individual Biomedical Research Award. This work was also supported by the National Institute for Neurological Disorders and Stroke (K02 NS112600), including support through the Center Without Walls on ion channel function in epilepsy (“Channelopathy-associated Research Center”, U54 NS108874), the Eunice Kennedy Shriver National Institute of Child Health and Human Development through the Intellectual and Developmental Disabilities Research Center (IDDRC) at Children’s Hospital of Philadelphia and the University of Pennsylvania (U54 HD086984), and by intramural funds of the Children’s Hospital of Philadelphia through the Epilepsy NeuroGenetics Initiative (ENGIN). Research reported in this publication was also supported by the National Center for Advancing Translational Sciences of the National Institutes of Health under Award Number UL1TR001878. This project was also supported in part by the Institute for Translational Medicine and Therapeutics’ (ITMAT) Transdisciplinary Program in Translational Medicine and Therapeutics at the Perelman School of Medicine of the University of Pennsylvania.

## Notes

### Competing Interest Statement

The authors have declared no competing interest.

### Author Declarations

The study was approved by the local ethical committees or followed other local guidelines as following: The ethics committee of Region Sjælland, Denmark: SJ-91 Ethics Committee Antwerp University Hospital, Drie Eikenstraat 655, 2650 Edegem, Belgium Ethical Committees of the Medical Faculty of the University of Munich Ethics Committee at the Research Centre for Medical Genetics, Moscow, Russian Republic Ethics committee of the University of Leipzig (ethical approval 224/16-ek, 402/16-ek)

## References

1. Veeramah KR, O’Brien JE, Meisler MH, et al. De novo pathogenic SCN8A mutation identified by whole-genome sequencing of a family quartet affected by infantile epileptic encephalopathy and SUDEP. American journal of human genetics 2012;90:502–510.

2. Anand G, Collett-White F, Orsini A, et al. Autosomal dominant SCN8A mutation with an unusually mild phenotype. European journal of paediatric neurology : EJPN : official journal of the European Paediatric Neurology Society 2016;20:761–765.

3. Gardella E, Becker F, Moller RS, et al. Benign infantile seizures and paroxysmal dyskinesia caused by an SCN8A mutation. Annals of neurology 2016;79:428–436.

4. Han JY, Jang JH, Lee IG, Shin S, Park J. A Novel Inherited Mutation of SCN8A in a Korean Family with Benign Familial Infantile Epilepsy Using Diagnostic Exome Sequencing. Annals of clinical and laboratory science 2017;47:747–753.

5. Johannesen KM, Gardella E, Encinas AC, et al. The spectrum of intermediate SCN8A-related epilepsy. Epilepsia 2019;60:830–844.

6. Estacion M, O’Brien JE, Conravey A, et al. A novel de novo mutation of SCN8A (Nav1.6) with enhanced channel activation in a child with epileptic encephalopathy. Neurobiology of disease 2014;69:117–123.

7. Ohba C, Kato M, Takahashi S, et al. Early onset epileptic encephalopathy caused by de novo SCN8A mutations. Epilepsia 2014;55:994–1000.

8. Vaher U, Noukas M, Nikopensius T, et al. De novo SCN8A mutation identified by whole-exome sequencing in a boy with neonatal epileptic encephalopathy, multiple congenital anomalies, and movement disorders. Journal of child neurology 2014;29:Np202–206.

9. Fung LW, Kwok SL, Tsui KW. SCN8A mutations in Chinese children with early onset epilepsy and intellectual disability. Epilepsia 2015;56:1319–1320.

10. Larsen J, Carvill GL, Gardella E, et al. The phenotypic spectrum of SCN8A encephalopathy. Neurology 2015;84:480–489.

11. de Kovel CG, Meisler MH, Brilstra EH, et al. Characterization of a de novo SCN8A mutation in a patient with epileptic encephalopathy. Epilepsy research 2014;108:1511–1518.

12. Rolvien T, Butscheidt S, Jeschke A, et al. Severe bone loss and multiple fractures in SCN8A-related epileptic encephalopathy. Bone 2017;103:136–143.

13. Wang J, Gao H, Bao X, et al. SCN8A mutations in Chinese patients with early onset epileptic encephalopathy and benign infantile seizures. BMC medical genetics 2017;18:104.

14. Gardella E, Marini C, Trivisano M, et al. The phenotype of SCN8A developmental and epileptic encephalopathy. Neurology 2018;91:e1112–e1124.

15. Johannesen KM, Gardella E, Scheffer I, et al. Early mortality in SCN8A-related epilepsies. Epilepsy research 2018;143:79–81.

16. Trudeau MM, Dalton JC, Day JW, Ranum LP, Meisler MH. Heterozygosity for a protein truncation mutation of sodium channel SCN8A in a patient with cerebellar atrophy, ataxia, and mental retardation. Journal of medical genetics 2006;43:527–530.

17. Blanchard MG, Willemsen MH, Walker JB, et al. De novo gain-of-function and loss-of-function mutations of SCN8A in patients with intellectual disabilities and epilepsy. Journal of medical genetics 2015;52:330–337.

18. Wagnon JL, Barker BS, Ottolini M, et al. Loss-of-function variants of SCN8A in intellectual disability without seizures. Neurology Genetics 2017;3:e170.

19. Liu Y, Schubert J, Sonnenberg L, et al. Neuronal mechanisms of mutations in SCN8A causing epilepsy or intellectual disability. Brain : a journal of neurology 2019.

20. Bunton-Stasyshyn RKA, Wagnon JL, Wengert ER, et al. Prominent role of forebrain excitatory neurons in SCN8A encephalopathy. 2019.

21. Wagnon JL, Mencacci NE, Barker BS, et al. Partial loss-of-function of sodium channel SCN8A in familial isolated myoclonus. Human mutation 2018.

22. Boerma RS, Braun KP, van de Broek MP, et al. Remarkable Phenytoin Sensitivity in 4 Children with SCN8A-related Epilepsy: A Molecular Neuropharmacological Approach. Neurotherapeutics : the journal of the American Society for Experimental NeuroTherapeutics 2016;13:192–197.

23. Sobreira N, Schiettecatte F, Valle D, Hamosh A. GeneMatcher: a matching tool for connecting investigators with an interest in the same gene. Human mutation 2015;36:928–930.

24. Scheffer IE, Berkovic S, Capovilla G, et al. ILAE classification of the epilepsies: Position paper of the ILAE Commission for Classification and Terminology. Epilepsia 2017;58:512–521.

25. Fisher RS, Cross JH, French JA, et al. Operational classification of seizure types by the International League Against Epilepsy: Position Paper of the ILAE Commission for Classification and Terminology. Epilepsia 2017;58:522–530.

26. Richards S, Aziz N, Bale S, et al. Standards and guidelines for the interpretation of sequence variants: a joint consensus recommendation of the American College of Medical Genetics and Genomics and the Association for Molecular Pathology. Genetics in medicine : official journal of the American College of Medical Genetics 2015;17:405–424.

27. Denis J, Villeneuve N, Cacciagli P, et al. Clinical study of 19 patients with SCN8A-related epilepsy: Two modes of onset regarding EEG and seizures. Epilepsia 2019;60:845–856.

28. Epi25. Ultra-Rare Genetic Variation in the Epilepsies: A Whole-Exome Sequencing Study of 17,606 Individuals. American journal of human genetics 2019;105:267–282.

29. Wagnon JL, Meisler MH. Recurrent and Non-Recurrent Mutations of SCN8A in Epileptic Encephalopathy. Frontiers in neurology 2015;6:104.

30. Pan Y, Cummins TR. Distinct functional alterations in SCN8A epilepsy mutant channels. The Journal of physiology 2020;598:381–401.

31. Schreiber JM, Tochen L, Brown M, et al. A multi-disciplinary clinic for SCN8A-related epilepsy. Epilepsy research 2020;159:106261.

32. Epifanio R, Zanotta N, Giorda R, Bardoni A, Zucca C. Novel epilepsy phenotype associated to a known SCN8A mutation. Seizure 2019;67:15–17.

33. Ranza E, Z’Graggen W, Lidgren M, et al. SCN8A heterozygous variants are associated with anoxic-epileptic seizures. American journal of medical genetics Part A 2020;182:1209–1216.

34. Wengert ER, Tronhjem CE, Wagnon JL, et al. Biallelic inherited SCN8A variants, a rare cause of SCN8A-related developmental and epileptic encephalopathy.

35. Lauxmann S, Verbeek NE, Liu Y, et al. Relationship of electrophysiological dysfunction and clinical severity in SCN2A-related epilepsies. Human mutation 2018;39:1942–1956.

36. Kohrman DC, Smith MR, Goldin AL, Harris J, Meisler MH. A missense mutation in the sodium channel Scn8a is responsible for cerebellar ataxia in the mouse mutant jolting. The Journal of neuroscience : the official journal of the Society for Neuroscience 1996;16:5993–5999.

37. Smith MR, Goldin AL. A mutation that causes ataxia shifts the voltage-dependence of the Scn8a sodium channel. Neuroreport 1999;10:3027–3031.

38. Dick DJ, Boakes RJ, Harris JB. A cerebellar abnormality in the mouse with motor end-plate disease. Neuropathology and applied neurobiology 1985;11:141–147.

39. Harris JB, Boakes RJ, Court JA. Physiological and biochemical studies on the cerebellar cortex of the murine mutants «jolting« and «motor end-plate disease«. Journal of the neurological sciences 1992;110:186–194.

40. Papale LA, Beyer B, Jones JM, et al. Heterozygous mutations of the voltage-gated sodium channel SCN8A are associated with spike-wave discharges and absence epilepsy in mice. Human molecular genetics 2009;18:1633–1641.

41. Makinson CD, Tanaka BS, Sorokin JM, et al. Regulation of Thalamic and Cortical Network Synchrony by Scn8a. Neuron 2017;93:1165-1179.e1166.

42. Wolff M, Johannesen KM, Hedrich UBS, et al. Genetic and phenotypic heterogeneity suggest therapeutic implications in SCN2A-related disorders. Brain : a journal of neurology 2017;140:1316–1336.

43. Liao Y, Deprez L, Maljevic S, et al. Molecular correlates of age-dependent seizures in an inherited neonatal-infantile epilepsy. Brain : a journal of neurology 2010;133:1403–1414.

44. Brunklaus A, Du J, Steckler F, et al. Biological concepts in human sodium channel epilepsies and their relevance in clinical practice. Epilepsia 2020;61:387–399.

45. Schwarz N, Bast T, Gaily E, et al. Clinical and genetic spectrum of <em>SCN2A</em>-associated episodic ataxia. European Journal of Paediatric Neurology 2019.

46. Ogiwara I, Miyamoto H, Tatsukawa T, et al. Nav1.2 haplodeficiency in excitatory neurons causes absence-like seizures in mice. Communications biology 2018;1.

47. Vacher H, Mohapatra DP, Trimmer JS. Localization and targeting of voltage-dependent ion channels in mammalian central neurons. Physiological reviews 2008;88:1407–1447.

48. Yu FH, Mantegazza M, Westenbroek RE, et al. Reduced sodium current in GABAergic interneurons in a mouse model of severe myoclonic epilepsy in infancy. Nature neuroscience 2006;9:1142–1149.

49. Ogiwara I, Miyamoto H, Morita N, et al. Nav1.1 localizes to axons of parvalbumin-positive inhibitory interneurons: a circuit basis for epileptic seizures in mice carrying an Scn1a gene mutation. The Journal of neuroscience : the official journal of the Society for Neuroscience 2007;27:5903–5914.

50. Bayat A, Hjalgrim H, Moller RS. The incidence of SCN1A-related Dravet syndrome in Denmark is 1:22,000: a population-based study from 2004 to 2009. Epilepsia 2015;56:e36–39.

51. Werling DM, Brand H, An JY, et al. An analytical framework for whole-genome sequence association studies and its implications for autism spectrum disorder. Nature genetics 2018;50:727–736.

52. Baker EM, Thompson CH, Hawkins NA, et al. The novel sodium channel modulator GS-458967 (GS967) is an effective treatment in a mouse model of SCN8A encephalopathy. Epilepsia 2018.

53. Lenk GM, Jafar-Nejad P, Hill SF, et al. Scn8a Antisense Oligonucleotide Is Protective in Mouse Models of SCN8A Encephalopathy and Dravet Syndrome. Annals of neurology 2020;87:339–346.

